# Extracellular vesicles as a liquid biopsy for amyotrophic lateral sclerosis: a systematic review and meta-analysis

**DOI:** 10.64898/2025.12.09.25341585

**Authors:** Magdalena M. Bolsinger, Nandhana Vivek, Javraj Singh, Ashrit Challa, Farbod Khorrami, Amanda Zhu, Trent Rothell, Samuel Wang, Nathan Robbins, Leony Fenwick, Grant Ruttenberg, Aleksander Bogoniewski, Hash Brown Taha

**Author notes:** **Corresponding Author:** Hash Brown Taha. contributed equally to this work (Javraj Singh, Ashrit Challa).

## Abstract

**Background:** Definitive diagnosis of amyotrophic lateral sclerosis (ALS) is only possible through a postmortem examination. Extracellular vesicles (EVs) have emerged as promising minimally invasive biomarkers for ALS, but studies vary widely in methodology and reproducibility. We conducted a systematic review and meta-analysis to evaluate the diagnostic potential of EV-associated proteins and RNAs in ALS.

**Methods:** Following PRISMA guidelines, we searched PubMed and EMBASE from inception to October 15, 2025. Thirty-nine studies met inclusion criteria. Random-effects models were used for continuous outcomes, and diagnostic accuracy was assessed using hierarchical summary ROC and bivariate random-effects models. Publication bias was evaluated using Begg, Egger, and funnel plots.

**Results:** EV-associated TDP-43 was the most frequently studied protein. Meta-analysis of five studies showed a moderate but non-significant increase in ALS vs. controls (SMD = 1.30) with high heterogeneity (I² = 97.8%). Sixteen studies assessing EV-RNA biomarkers showed minimal overlap and limited independent replication. Diagnostic accuracy meta-analysis across 11 studies yielded moderate performance (AUC = 0.839). No publication bias was found across both meta-analyses.

**Conclusions:** EV biomarkers for ALS show biological promise but are limited by methodological variability and insufficient replication. Standardized protocols, transparent data sharing, and independent validation are needed.

## Introduction

Amyotrophic lateral sclerosis (ALS) is a fatal and complex neurodegenerative syndrome marked by the progressive degeneration of both upper and lower motor neurons. This neuronal loss leads to denervation of skeletal muscles and ultimately to paralysis. Neuropathologically, cytoplasmic aggregation of TAR DNA-binding protein 43 (TDP-43) in degenerating motor neurons represents the defining hallmark of ALS, present in approximately 97% of persons with ALS^1, 2^. Although the precise etiology remains elusive, converging evidence indicates that ALS arises from a complex interplay of genetic susceptibility and environmental exposures^3^. While familial ALS with identified causative mutations only accounts for 5-10% of patients, most cases are sporadic^4^.

ALS diagnosis relies primarily on clinical judgment, based on the recognition of characteristic motor features and exclusion of mimicking disorders. Despite refined consensus criteria, diagnostic confirmation remains challenging, particularly early in the disease course complicated by high clinical symptom heterogeneity. Consequently, delays between symptom onset and diagnosis are common, often extending over a year^5, 6^. At present, no effective disease-modifying therapy is available for ALS, and median survival following diagnosis is approximately three years^7^. Given the diagnostic delay and limited efficacy of current treatments, the development of validated, non-invasive biomarkers is an urgent priority. Such biomarkers could enable earlier diagnosis, offer quantitative measures of disease progression (prognosis) and stratify patients for clinical trials to evaluate treatment outcomes (predictive).

Numerous approaches have been explored to improve the diagnosis of ALS including neurofilament light chain (NfL) and phosphorylated NfL in cerebrospinal fluid and blood, neuroimaging techniques such as MRI and diffusion tensor imaging, electrophysiological testing, inflammatory and metabolic biomarkers, and genetic screening for common ALS-associated mutations^8–13^. Yet, a definitive diagnosis remains possible only through postmortem neuropathological confirmation characterized by motor neuron loss and TDP-43-positive inclusions.

Extracellular vesicles (EVs) are a heterogeneous population of small, membrane-bound vesicles, including ectosomes and exosomes^14^. Encapsulated by a phospholipid bilayer, EVs carry diverse cargos of proteins, lipids, carbohydrates, and nucleic acids and function in intercellular communication as well as cellular waste disposal^15^. Notably, pathogenic proteins associated with ALS and other neurodegenerative diseases have been identified from EVs^16^. Because EVs may traverse the blood–brain barrier and reflect the molecular state of their cells of origin, both general EVs and so-called speculative CNS-enriched EVs have been proposed as minimally invasive biomarker sources for neurological disorders^17–20^. In our recent large-scale meta-analyses of parkinsonian disorders^18, 19^ and dementia^20^, we found that biomarkers derived from general EVs demonstrated superior diagnostic accuracy compared with speculative CNS-enriched EVs, with RNA-based EV biomarkers showing the strongest overall performance.

ALS-specific EVs for biomarker development and preclinical diagnosis have been investigated in recent years^21^. However, with differences in cohorts, biofluids, EV extraction methods, and analytical platforms individual studies often yield variable and underpowered results. To allow clinical translation of identified biomarkers in basic research to clinical use, a comprehensive understanding of the to date identified EV-associated biomarkers is needed. As such, we conducted a systematic review and meta-analysis of published studies investigating EV-associated molecular signatures in ALS.

## Methods

We conducted a systematic review, meta-analysis, and meta-regression following the guidelines of the Preferred Reporting Items for Systematic Reviews and Meta-Analyses (PRISMA). The study used only anonymized, previously published data and did not involve the collection of personal information or any direct research on human participants; therefore, ethical approval was not required. The study protocol was not registered.

### Data sources and search strategy

We performed a thorough search for relevant articles by using specific search terms related to ALS and related disorders. The search was conducted in two databases (PUBMED and EMBASE) and covered articles published from the inception of the databases until October 15th, 2025. Three independent researchers (HBT, NV & JS) screened all the titles, abstracts, and full manuscripts to select articles that met the eligibility criteria. By hand, we reviewed the reference lists of eligible studies and searched Google scholar for articles using EVs as a liquid biopsy for ALS. Any discrepancies in article selection were resolved through discussion. The complete search strategy can be found in **Table S1.**

### Eligibility criteria

To be included in the systematic review (qualitative), the eligible studies had to use EVs as a liquid biopsy for ALS in humans. We excluded studies that used animals or in vitro models (cell culture lines, primary cells and iPSCs) and studies that did not include the specified diseases. For studies that included longitudinal measurements or treatment interventions, we only considered the baseline assessments. Additionally, we contacted all authors to obtain other relevant information (see **Table 1** for detailed information).

To be included in the quantitative random-effects meta-analysis, studies were required to report a biomarker with mean and standard deviation and the corresponding group sizes. For studies that did not provide mean and standard deviation, we first searched for publicly available datasets and directly contacted the authors to request the raw values. If no dataset was available, we examined the figures to determine whether individual data points were overlaid; when present, we extracted those values using WebPlotDigitizer v3.4 (https://web.eecs.utk.edu/∼dcostine/personal/PowerDeviceLib/DigiTest/index.html) to obtain the individual values and calculate the mean ± SD. From one study^22^, we estimated the mean and SD by transforming the median (Q1 + median + Q3)/(3) and IQR (Q3-Q1)/(1.35) as established previously^23^.

To be included in the quantitative diagnostic accuracy meta-analysis, studies needed to have conducted an ROC analysis and reported the AUC, sensitivity, and specificity, and either provided the underlying dataset or included figures with overlaid individual values from which we could estimate these metrics using WebPlotDigitizer v3.4. We employed this framework to conduct a binomial logistic regression and obtain the sensitivity and specificity at the cutoff that optimizes Youden’s index^24^.

### Data extraction

Data from eligible studies were extracted by at least two independent researchers. Authors checked the database for accuracy and completeness. Authors from the respective studies were contacted to obtain any missing information listed in Table 1. For quantitative outcomes, means and standard deviations were taken directly from the original articles. When numerical values were not reported but individual data points were shown graphically, missing data were obtained with the open-source tool WebPlotDigitizer (https://automeris.io/). The extracted values were then validated for accuracy using GraphPad Prism (version 8.0).

### Risk of bias assessment

We did not perform a formal quality or risk of bias assessment using standardized tools due to the heterogeneous and complex nature of the included studies. Instead, we ensured methodological rigor and data reliability by carefully screening study eligibility, verifying extracted data, and directly contacting the corresponding authors to obtain missing information or clarify uncertainties.

### Data synthesis and statistics

Random□effects meta□analyses were performed in R software (version 2024.12.0+467) using the *metafor* package. An inverse variance weighting model was applied, and pooled standardized mean differences (SMD) with 95% confidence intervals were calculated based on Cohen’s d using the following formula:

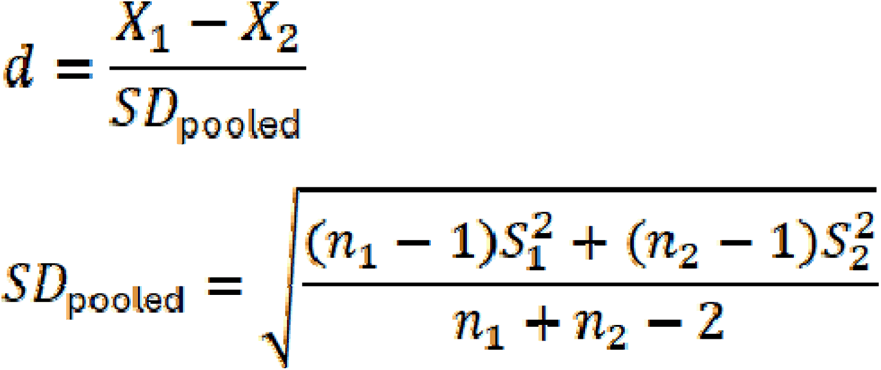

We performed diagnostic accuracy meta-analyses using the hierarchical summary ROC (HSROC) model or the bivariate random effects meta-analysis (BRMA) model, which are mathematically equivalent when no covariates are included^25^. These models were conducted using the *metadta*^26^ command in Stata, which has been extensively validated in comparison to other packages^27^. Model selection (structured vs unstructured covariance) was based on the lowest Akaike and Bayesian information criteria (AIC, BIC), with AIC used to break ties. Accuracy was interpreted from HSROC curves, where estimates closer to the upper left quadrant indicate higher performance. Heterogeneity was quantified using the I² statistic of Zhou and Dendukuri^28^.

Publication bias^29^ was evaluated using Begg’s rank correlation, Egger’s and Deek’s regression tests and funnel plots. Studies with sensitivity and specificity = 1 were excluded from the diagnostic accuracy publication bias assessment. All publication bias analyses were performed in R software (version 2024.12.0+467).

### Terminology

We use the term general EVs to refer to EVs isolated using bulk isolation approaches that do not involve targeted immunoprecipitation for speculative CNS markers, the term speculative CNS-enriched EVs to refer to EV enriched using speculative markers such as L1CAM for neuronal EVs (nEVs) or ACSA for astrocytic EVs, and CNS EVs to refer to EVs directly isolated from CNS tissue sources including brain and spinal cord. This is for at least three key reasons. The enrichment markers used are not specific to brain tissue and can also exist freely in circulation or on diverse peripheral cells. In addition, because EVs are continuously taken up and recycled by numerous cells throughout the body, any original signal they may have carried from the brain is likely diluted or altered. Lastly, no study has confirmed or proven that these EVs do indeed originate from the CNS. Finally, CNS EVs refer to EVs isolated directly from CNS tissue.

## Results

### Cohort description

Our systematic reviews encompassed 39 studies^22, 30–67^, of which 15 were included in the meta-analyses (**Figure 1**). The study population comprised 1,142 persons with ALS, 847 controls, 244 persons with frontotemporal dementia (FTD), 66 with ALS-FTD, 118 with other motor neuron diseases, 87 with Parkinson’s disease (PD), 12 with Alzheimer’s disease (AD), 207 with progressive supranuclear palsy (PSP), 3 with idiopathic normal pressure hydrocephalus, 3 with other amyloidogenic disorders, and 9 with unspecified diseases (see **Table 1** for full details). 27 studies used general EVs. 8 studies used speculative CNS-enriched EVs using neural or glial markers (e.g., L1CAM, GLAST, ACSA-1, GAP43, NLGN3), 1 study used general EVs and speculative CNS-enriched EVs, 2 studies used CNS EVs, including whole brain and motor cortex samples, and 1study used general and CNS EVs from brain and spinal cord. Plasma was used in 17, CSF in 8 and serum in 9 studies. Defibrinated plasma was used in 3 studies, and saliva in 1study. In addition to peripheral biofluids, 3 studies examined CNS tissue sources, including temporal cortex, frontal cortex, motor cortex, and spinal cord. Across the included studies, ultracentrifugation (UC) was the most used EV isolation method (11 studies), including sucrose gradient-UC (2 studies). Polymer-based precipitation was used in 10 studies, primarily through ExoQuick. Membrane-affinity based approaches such as exoRNeasy or exoEasy were applied in 5 studies, while combinations of SEC, UF, and UC were reported in 4 studies. Less frequently used approaches included affinity capture (Vn96 peptide) in 2 studies, EXOBead, nickel-based isolation, ExoSORT™ (citation), and flow cytometry. In terms of EV characterization, 19 studies report TEM, 19 studies report NTA or comparable particle sizing/counting, and 21 studies include western blots (WBs) for EV markers (such as CD63, CD81, TSG101, syntenin or flotillin) and negative markers (such as calnexin, GM130 or mitochondrial/ER proteins), whereas the remaining 12 studies either provide minimal EV phenotyping or only indirect confirmation.

**Figure 1.**
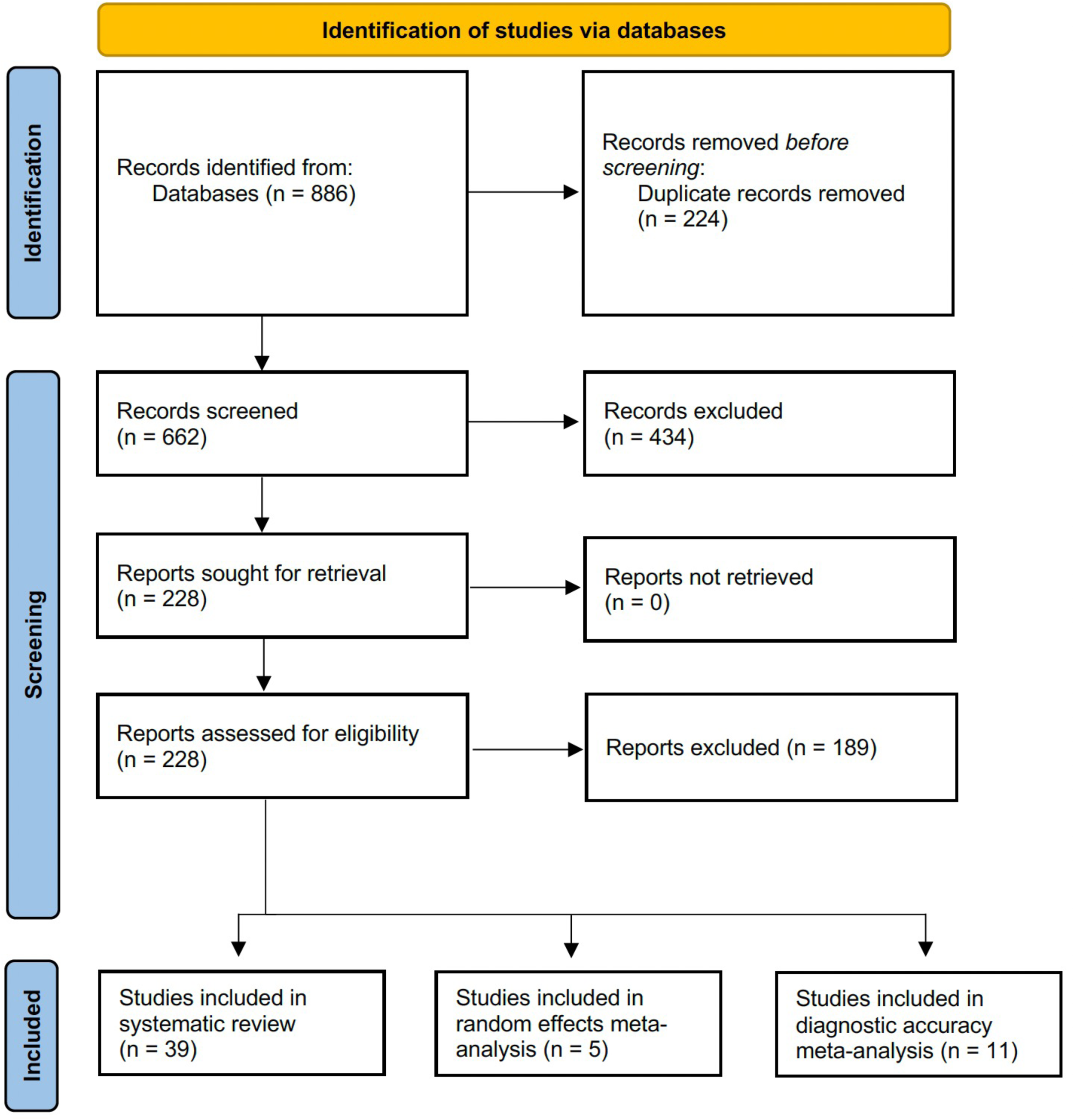
PRISMA flow-diagram.

Protein-focused studies quantified specific cargo such as total or phosphorylated TDP 43, C □terminal TDP □43 fragments, SOD1, FUS, HSP90, pNFH, HERV □K, total tau, 3R/4R □tau, IL □6 and tetraspanins using WB, ELISA, single molecule array or mass spectrometry. Sixteen studies are RNA-centered and analyze miRNAs, mRNAs, lncRNAs or unannotated RNAs, typically testing panels of 10 to 30 prespecified miRNAs, more than 80 RNAs in multiplex platforms, or discovering dozens to more than 100 dysregulated RNAs per EV fraction via small RNA □seq, total RNA □seq, microarrays, qPCR or ddPCR.

### Proteins

#### TDP-43, SOD1 and FUS

TDP-43, SOD1, and FUS are key proteins implicated in the pathogenesis of ALS with abnormal aggregation, mislocalization, or mutation of these proteins contributing to motor neuron dysfunction and degeneration^68^. TDP-43 pathology is present in the vast majority of ALS cases and is considered a defining neuropathological hallmark, while mutations in SOD1 and FUS represent established genetic causes of familial and sporadic ALS^68^. In our systematic review, 9 studies have quantified TDP-43 in general EVs isolated from CSF^30, 32^, plasma^22, 35, 44, 49^, serum speculative CNS-enriched EVs^56^ and brain tissue^33, 41^.

In CSF, 2 studies suggested that TDP-43 levels may be higher in those with FTD pathology as compared to ALS alone. The first study detected TDP-43 derived from general EVs of individuals with ALS using WB, but subsequent mass spectrometry analysis, with normalization to flotillin-1, did not show a statistically significant difference between ALS and controls although TDP43 trended toward higher levels in FTD^30^. In contrast, the second study^32^ demonstrated that various TDP-43 species analyzed via WB (full-length TDP-43, TDP-35, and TDP-25) were higher in ALS-FTD vs. controls. Both studies suggest that CSF EV-associated TDP-43 and its cleavage products may be more prominently increased in ALS-FTD than in ALS alone.

A recent seminal study^22^ with two independent large cohorts measured TDP-43 in plasma general EVs using single molecule array. Across cohorts, plasma EV TDP-43 was consistently highest in ALS. Specifically, in the first cohort, (DESCRIBE SUBCOHORT 2), TDP43 levels in ALS were distinctly higher when compared to controls, behavioral variant FTD (bvFTD), and PSP. Furthermore, EV-associated TDP-43 distinguished ALS from controls, PSP, and bvFTD with AUCs around 0.91-0.99. Cases with confirmed TDP43 pathology (genetic or neuropathologic) had markedly elevated EV TDP-43 compared with controls, PSP-type tau pathology, MAPT mutation carriers, and non-TDP-43/non-tau cases. Non-TDP-43/non-tau cases (SOD1, FUS, CHCHD10) had low TDP-43, similar to controls and PSP. In the second cohort (Sant Pau), the pattern was replicated: ALS and ALS-FTD had the highest plasma EV TDP-43 levels exceeding those of controls and PSP, and a subset of bvFTD cases also showed elevated EV TDP43, clustering with ALS. In both cohorts, EV TDP-43 outperformed plasma NfL, a widely used marker of neurodegeneration^69^, in distinguishing bvFTD from controls and PSP, as well as ALS/ALS-FTD from controls and PSP in cohorts where NfL data were available. The same pattern was observed in the second cohort where genetic cases associated with TDP43 pathology (C9orf72, GRN, VCP, TBK1, etc.,.) showed high plasma EV TDP43 levels, whereas genetically confirmed non-TDP43/non-tau cases (SOD1, FUS) had low levels, comparable to controls and PSP. Using cohort-specific cut-offs (for example >13.87 or >17.85 pg/ml), EV TDP43 accurately identified TDP43 pathology with high sensitivity and very high specificity in both cohorts and robustly detected sporadic ALS and ALS-FTD vs. controls and PSP. Additionally, EV plasma TDP43 behaved like a severity marker correlating with disease severity and plasma NfL in both ALS and bvFTD.

These findings were previously supported by a study^44^ of 18 people with ALS, where plasma EV TDP43 ratios were measured at baseline and at 1-, 3-, 6-, and 12-month follow-up visits using flow cytometry. EV TDP-43 was significantly higher at 3 and 6 months compared with baseline, with a borderline increase at 12 months, suggesting progressive accumulation of EV-associated TDP-43 over time. When patients were stratified by clinical course, increases in plasma EV TDP-43 occurred earlier in those who later demonstrated more rapid functional decline. Additionally, another study measured plasma EV TDP-43 levels in putative larger-sized and smaller-sized EVs using WB and found that total TDP-43 and phosphorylated TDP-43 were higher in putative larger-sized EVs from persons with ALS compared to controls, while no significant differences were observed in smaller-sized EVs. Importantly, the ranges of TDP-43 levels in ALS and control samples overlapped substantially, indicating that although putative larger-sized EVs showed an overall increase in TDP-43, these levels alone were insufficient to reliably distinguish individuals with ALS from controls.

It is worth noting that one study reported TDP-43 detection in general plasma EVs by WB was challenging, due in part to interference from abundant plasma proteins such as immunoglobulins and albumin when using antibodies against the N- or C-terminus. Although a phosphorylated TDP-43 antibody produced a 45 kDa doublet in both ALS and control samples, immunogold TEM failed to confirm intravesicular localization, suggesting the signal was not truly EV-associated. This raises concerns about the specificity and reproducibility of measuring TDP-43 in plasma general EVs^49^.

From one study^56^ measuring TDP43 in speculative CNS-enriched nEVs using ELISA, TDP43 levels were higher in people with ALS compared to controls at baseline. In the longitudinal treatment arm (treated with a combination of ciprofloxacin and celecoxib), EV-associated TDP43 levels decreased over time with therapy, while conventional plasma markers such as NfL and pNfH remained largely unchanged. In a postmortem study^34^, CNS EVs isolated from the temporal cortex of 3 individuals with sporadic ALS contained significantly higher levels of TDP43, including full-length and C-terminal fragments, compared with EVs isolated from individuals with other neurologic disorders (1 multiple system atrophy, 1 familial amyloid polyneuropathy, and 1 chronic inflammatory demyelinating polyneuropathy). In a postmortem study^33^, general EVs isolated from the motor cortex of individuals with ALS were significantly enriched in the 28 kDa C-terminal fragment of TDP-43 compared with neurological controls, indicating that ALS brain-derived EVs carry higher levels of pathogenic TDP-43 species. This may support a direct link between CNS TDP43 pathology packaging into EV cargo in ALS^41^.

Studies measuring SOD1 and FUS1 in EVs are scarce; however, one study^35^ reported that SOD1 and FUS1 were higher in plasma putative larger-sized EVs from persons with ALS compared to controls, whereas no clear disease-related differences were seen in putative smaller-sized EVs.

#### TDP43 random-effects meta-analysis

The random effects meta-analysis of five studies^22, 30, 32, 35, 56^ showed an overall increase in EV associated TDP-43 in ALS compared with controls (SMD = 1.30; **Figure 2A**), though this did not reach statistical significance (p = 0.068), largely due to extremely high heterogeneity (I² = 97.8%). One study^36^ reporting low TDP-43 levels may have contributed disproportionately to this heterogeneity. We removed this outlier in a sensitivity analysis, and the pooled effect increased (SMD = 1.87) and almost approached statistical significance (p = 0.052). We further assessed publication bias, and Begg’s correlation (Kendall’s tau = 0.14, p = 0.77), Egger’s regression test (t = 0.28, p = 0.79) and visual inspection of funnel plot suggest very minimal bias, although one study with an extreme effect size increased apparent scatter (**Figure 2B**). These results suggest that more studies are needed to draw definitive conclusions on whether EV-associated TDP-43 is higher in ALS vs. controls.

**Figure 2.**
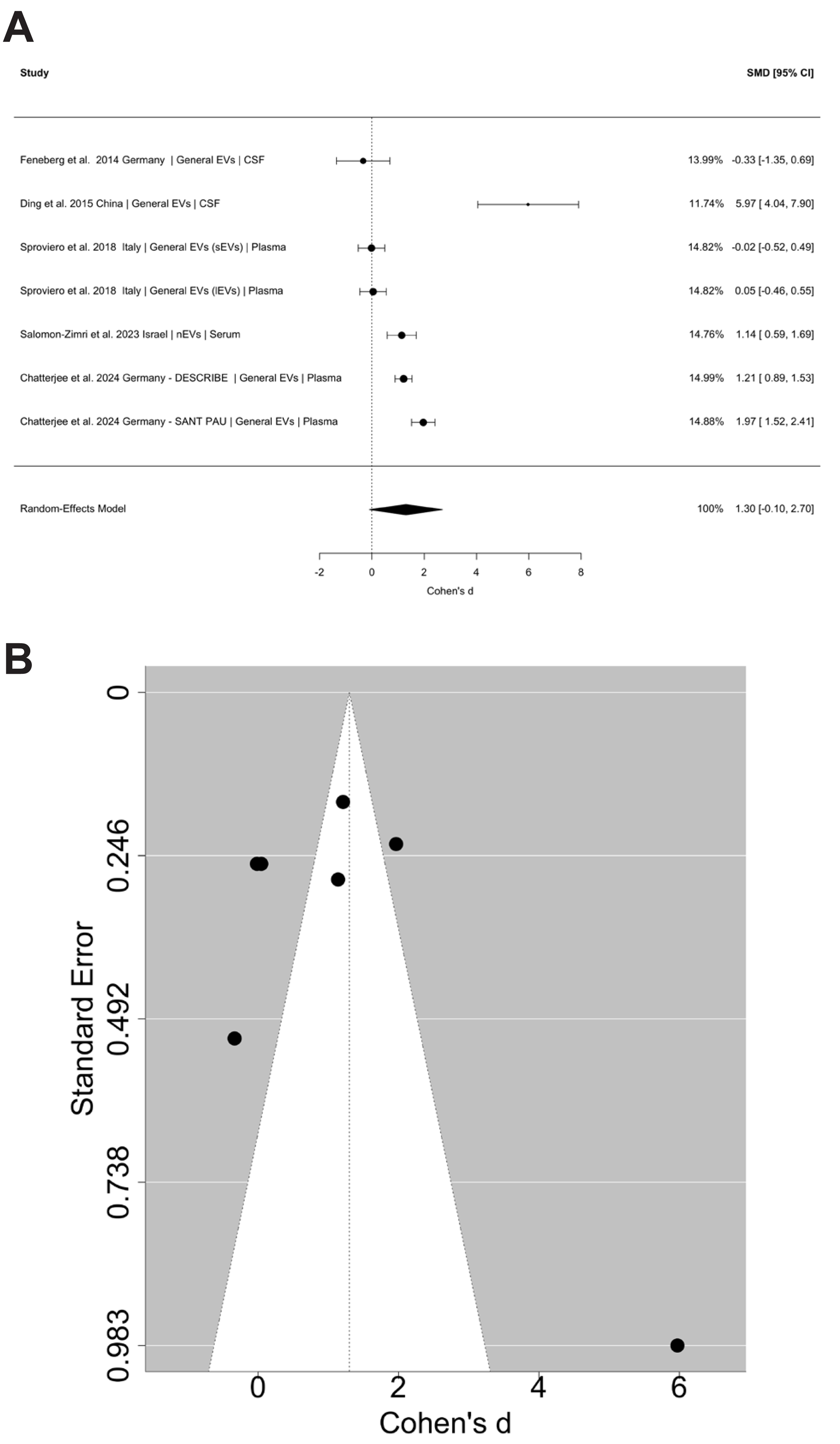
Meta-analysis of EV-associated TDP-43 levels in amyotrophic lateral sclerosis (ALS). **(A)** Forest plot summarizing standardized mean differences (SMDs; Cohen’s d) for EV-associated TDP-43 levels comparing ALS cases with controls across 5 independent studies. Study labels include country, EV subtype, and biofluid source. Squares represent individual study effect sizes scaled by study weight, with horizontal lines indicating 95% confidence intervals. The diamond represents the pooled random-effects estimate, showing a trend toward higher EV-associated TDP-43 in ALS, although non-significant. sEVs = small extracellular vesicles; lEVs = large extracellular vesicles. **(B)** Funnel plot assessing publication bias for the included studies. Each point represents an individual study plotted by effect size (Cohen’s *d*) and standard error. Visual inspection shows no strong asymmetry, consistent with statistical tests indicating no evidence of publication bias. Though one study appears to have increased scattering.

#### Other biomarkers

Beyond TDP-43 pathology, several additional biomarkers derived from EVs in ALS have been explored, reflecting diverse biological processes that extend beyond protein aggregation alone, including inflammatory and immune-related signals (IL-6, GLAST), disruptions in protein homeostasis (HSP90), markers of axonal injury (pNFH), extracellular matrix-associated proteins (PRG-4), potential endogenous retroviral activity (Retrovirus), and biophysical molecular signatures (Raman spectroscopy).

Two studies provided evidence of inflammatory changes in EVs from persons with ALS. IL-6 levels were reported to be higher in speculative astrocytic EVs from people with ALS compared to controls and correlated with faster disease progression, particularly in early disease^38^, whereas speculative CNS-enriched GLAST positive EV counts were also increased in ALS but did not correlate with clinical severity measures, limiting their reliability^63^. Importantly, the absolute counts of leukocyte, endothelial and platelet derived EVs were similar between ALS and controls, suggesting that ALS may involve altered EV cargo rather than increased EV release from inflammatory cells^62^. Additionally, one study^49^ evaluated HSP90, PPIA, and pNFH in general plasma EVs, and found that HSP90 was significantly lower in plasma-derived EVs from people with ALS compared with controls and spinal and bulbar muscular atrophy and was similarly decreased in EVs from ALS mouse models, suggesting that lower EV-associated HSP90 may reflect ALS pathology across sporadic and genetic forms. In contrast, EV-associated PPIA levels did not differ between groups, indicating limited diagnostic value, while pNFH (another marker of neuronal injury similar to NfL) was markedly increased in plasma from people with ALS, particularly in those with faster progression, but could not be reliably detected from EVs, suggesting that pNFH may not meaningfully be packaged into EV cargo.

Some other notable biomarkers derived from EVs have been quantified in ALS, spanning protein, viral, and spectroscopic signatures. PRG-4 was found to be significantly enriched in general EVs from persons with ALS, particularly in those with preserved cognition, and this increase was confirmed by ELISA, suggesting a possible protective or compensatory association with cognitive status^60^. In contrast, HERV-K env measured in speculative nEVs did not differ overall between persons with motor neuron disease (MND; 23 out of 39 with ALS) and controls, but levels were higher in individuals with classical ALS compared to other MND phenotypes and tended to increase with longer disease duration and greater lower motor neuron involvement^42^. Finally, Raman spectroscopy revealed distinct biochemical signatures in general EV populations from ALS compared to controls, with larger EVs showing increased lipid-related peaks and reduced protein-related peaks, and smaller EVs showing more subtle lipid-related shifts. Multivariate analysis of the Raman spectra modestly discriminated ALS from controls, supporting EV biochemical composition as a potential complementary, though not definitive, biomarker of disease state^42^.

#### Proteomics

Ten studies^30, 31, 41, 45, 46, 54, 60, 63, 65, 67^ have used proteomics to evaluate biomarkers derived from EVs (see **Table S2** for a summary). Only one compared ALS to PD^31^, and the other discussed EV proteomics only in relation to the TDP-43/flotillin-1 ratio^30^.

A proteomic study^41^ using speculative CNS-enriched EVs isolated from the postmortem motor cortex of individuals with ALS identified twelve distinct proteins that were only found in ALS CNS EVs: CD177, CHMP4B, CSPG5, DYNC1I2, IGHV3-43, LBP, RPS29, S100A9, SAA1, SCAMP4, SCN2B, and SLC16A1, several of which are associated with inflammation (S100A9, SAA1, LBP), vesicle biogenesis (CHMP4B), cytoskeletal transport (DYNC1I2), or neuronal excitability (SCN2B). In addition, 16 proteins were differentially expressed between ALS and controls (VCAM1, STAU1, RRAS, PLSCR4, NT5E, ITGA5, HLA-A, GYPC, FXYD6, ENPP6, ENG, EHD1, DYNC1I1, DHX30, BST1, and AHNAK), reflecting alterations in immune signaling, RNA metabolism, membrane remodeling, and cytoskeletal function in ALS.

A small proteomic study that quantified CSF EVs from 3 persons with ALS and 3 persons with idiopathic normal pressure hydrocephalus identified several differentially expressed proteins, though findings are limited by the small sample size^46^. Three proteins were increased in ALS EVs (NOC2L, PDCD6IP, and VCAN), while several others were decreased, including SERPINA3, PTPRZ1, C1QC, CCDC19, MYL6B, MARCO, FCGBP, FOLR1, RELN, CFB, and CHMP4A. Another CSF EV proteomics study identified only one biomarker, bleomycin hydrolase (BLMH), which was significantly downregulated in ALS compared to controls^45^. These preliminary results point to possible EV associated changes in inflammatory, synaptic, and matrix related pathways in ALS, but none of these proteins have been reported in other proteomic studies or validated using targeted approaches. Another proteomic screen of plasma derived EVs from 3 persons with ALS and 3 controls identified twenty top candidate proteins that were markedly enriched in ALS EVs^54^. These included several RNA binding and splicing related proteins (HNRNPD, HNRNPA0, HNRNPC, HNRNPA1, RBMX, PUF60, TCERG1, SF3B3, DEK, DDX5), cytoskeletal and actin related proteins (CORO1A, ACTG1, ACTG2, ARPC1B, IQGAP1), and proteins implicated in ALS pathology such as FUS and PADI4. Many of these proteins showed more than three-to-five-fold higher abundance in ALS EVs compared with controls, suggesting potential alterations in RNA processing, cytoskeletal regulation, and stress response pathways in circulating EVs from persons with ALS. A plasma EV proteomic study that included both a discovery and an independent validation cohort identified seven consistently altered proteins in ALS, all of which were related to platelet or immune pathway activity. These proteins were LBP, FGB, FGG, C9, FGA, PRG4, and VWF, highlighting a reproducible signature of coagulation and complement-associated EV cargo changes in ALS^60^.

A serum EV proteomics study comparing people with ALS to controls found a distinct pattern of protein distribution rather than broad shifts across the EV proteome^63^. EVs from persons with ALS were relatively enriched in TALDO1, PTRC, PIP4K2A, LGALS7, GP9, TMP3, FGB, P4HB and CD36, whereas EVs from controls contained higher levels of SERPING1, IGHA2, IGLC3, APOC1, APOB, F11, SLC4A1 and APMAP. Among these, IGHA2 was reported to separate both groups the most. They also performed an EV metabolomics analysis, which revealed clear group-specific metabolic signatures. EVs from people with ALS were enriched in sphingomyelin, 5-valerolactone, 2-OH-4-methylpentanoate, 3,4-di-OH-phenylacetate, whereas EVs from controls showed higher levels of phosphatidylcholine, L-carnitine, isocitrate, suberate, deoxycarnitine, 24-hydroxycholesterol, 2-methylbutyrylglycine, and 3-methylcrotonylglycine.

Two studies^53, 58^ applied proximity extension assay to EVs in ALS but did not identify robust disease associated signals. In CSF and CSF derived general EVs, dozens of proteins were detectable, but neither hierarchical clustering nor principal component analysis produced a clear separation between ALS and controls, and no consistently differentially expressed EV proteins emerged; only crude CSF showed partial separation driven by CHIT1 and myoglobin. In the second study^58^ analyzing saliva and saliva derived EVs also found no group separation on principal component analysis and no significantly altered proteins, with only weak trends toward lower ZNF428 in saliva EVs and higher IGLL1 in whole saliva from people with ALS. These suggest that proximity extension assays may not be a good method for finding differentially expressed biomarkers derived from EVs. Lastly, one study found increased levels of coronin-1a in plasma EVs of 3 persons with ALS vs. controls, but did not validate it using targeted-approaches in EVs^54^.

Lastly, a longitudinal serum EV proteomic study^65^ tracked individuals with SOD1 D90A ALS (plus two additional ALS mutation carriers) across disease stages and showed that EV protein profiles shift markedly with symptom onset and progression. Early disease was characterized by vascular and genomic stress signals, symptomatic conversion by strong immune and complement activation, and advanced disease by intensified inflammatory and phagocytic pathways. Importantly, FN1 and multiple FN1 isoforms increased consistently with clinical worsening, identifying FN1 as a potential EV-based progression marker.

### RNA

Both DNA and RNA are present in EVs and can serve as biomarkers, with EV cargo including mRNA, transfer RNA, circular RNAs and other noncoding RNAs. In our two previous meta-analyses on parkinsonian disorders^18, 19^ and dementia^20^, RNA-based EV biomarkers showed the highest diagnostic accuracy and overall performance compared with other biomarkers. In the context of ALS, 16 studies have quantified RNA levels derived from EVs, six^34, 43, 50, 55, 61, 66^ using qPCR to measure specific miRNAs, four^37, 47, 51, 52^ applying RNA sequencing or microarray approaches to identify broader panels of differentially expressed RNAs, and 6 using combined qPCR and RNA sequencing^36, 39, 40, 57, 59, 64^ (see **Table 1**).

The main conclusion from these studies is that the findings show minimal overlap, frequently originate from the same research groups, and lack independent replication, making it difficult to draw reliable or generalizable conclusions about RNA-based EV biomarkers in ALS currently. While similarity regarding pathway detection has been found across studies, only a few miRNAs were even validated via qPCRs in their own studies. This suggests that miRNAs as cargoes in EVs might be interesting tools to study the underlying pathophysiology but questions their actual use as reliable and reproducible diagnostic biomarkers for ALS, at least by using conventional laboratory tools like qPCR. Below we discuss a few of these studies.

One study reported the miRNA-27a-3p to be down regulated in EVs isolated from the serum of people with ALS^34^. The authors speculated that miRNA-27a-3p, a microRNA previously found to be implicated in neuronal stress responses, plays a role in mitigating the communication between muscle and bone in the context of disease, possibly reflecting dying muscle tissue in ALS. Another CSF EV study^43^ reported that miR-124-3p, a microRNA involved in neuronal differentiation and neuroinflammation, showed a trend toward higher levels in male persons with ALS and correlated strongly with worse disease severity (ALSFRS-R), although the group difference did not reach statistical significance. This relationship was not observed in female patients, and miR-124-3p levels were unrelated to age or body size, suggesting a possible sex-specific association between CSF EV miR-124-3p and ALS progression. A recent study analyzed plasma EV miRNAs in ALS using discovery and validation cohorts stratified by SOD1 and C9orf72 status^56^. Microarray screening followed by qPCR showed miR-34a-3p to be selectively reduced in SOD1-ALS and miR-1306-3p to be significantly downregulated in persons with genetic ALS with SOD1 or C9orf72 mutations. The miR-199a-3p and miR-501-3p were elevated in sporadic ALS compared to controls, an effect mainly observed in males. miR-501-3p was further enriched in persons with bulbar-onset disease compared with spinal-onset or classical Charcot phenotypes^701^, suggesting value for phenotypic discrimination. A machine-learning classifier using five miRNAs (miR-199a-3p, miR-30b-5p, miR-501-3p, miR-103a-2-5p, miR-181d-5p) achieved an AUC of 0.80 with 79% accuracy, supporting circulating, blood-derived EV-miRNAs as informative biomarkers that distinguish ALS from controls and capture genotype-specific signatures.

Additionally, a study using plasma EVs found altered expression of miR-449a and its predicted target ADAM10 in ALS compared to controls. Specifically, miR 449a was upregulated in EVs from persons with ALS, while ADAM10 showed an opposite, downregulated trend, supporting a reciprocal miRNA-silencing relationship consistent with neurodegenerative mechanisms^64^.

Several studies assessing miRNAs in speculative CNS-enriched EVs have reported differences between ALS and controls. However, multiple studies reporting these findings^40, 55, 61^ were produced by the same group, and the authors did not provide their datasets despite repeated requests. No independent group has replicated these miRNA results, and no other studies have examined these specific candidates in speculative nEVs, leaving their biomarker value uncertain. Additional studies have analyzed other biomarkers in speculative nEVs^37, 48, 56^ and astrocyte-enriched EVs^38, 62^, but none show overlap in identified biomarkers across groups. Importantly, speculative CNS-enriched EVs consistently show lower diagnostic performance and markedly greater publication bias compared with general EVs across multiple meta-analyses^18–20^

Ten studies have examined miRNA profiles within EVs using RNA sequencing technologies^36,37, 39, 40, 47, 51, 52, 57, 59, 64^.

In one such investigation of serum-derived EVs from individuals with ALS and controls, no miRNA species reached statistical significance in association with ALS pathology^50^. The authors contrasted their findings with two prior reports^37, 40^ each of which analyzed speculative nEVs isolated from plasma. These earlier studies described modest sets of differentially expressed miRNAs, 13 upregulated and 17 downregulated in one, and 5 upregulated and 3 downregulated in the other, yet no overlap emerged between them. Importantly, neither study applied correction for multiple testing, a methodological omission that substantially increases the likelihood of false-positive results^37, 40^. A recent study analyzed plasma EVs miRNAs in ALS using discovery and validation cohorts stratified by SOD1 and C9orf72 status^57^. Microarray screening followed by qPCR showed miR-34a-3p to be selectively reduced in SOD1-ALS and miR-1306-3p to be significantly downregulated in both persons with genetic ALS with both SOD1 or C9orf72 mutations, while miR-199a-3p and miR-501-3p were elevated in sporadic ALS. miR-501-3p was further enriched in persons with bulbar-onset disease compared with spinal-onset or classical Charcot phenotypes^70^, suggesting value for phenotypic discrimination. A machine-learning classifier using five miRNAs (miR-199a-3p, miR-30b-5p, miR-501-3p, miR-103a-2-5p, miR-181d-5p) achieved an AUC of 0.80 with 79% accuracy, supporting circulating, blood-derived EV-miRNAs as informative biomarkers that distinguish ALS from controls and capture genotype-specific signatures. Another group focused on the analysis of mRNAs isolated from CSF-derived EVs from 4 persons with ALS and 4 controls using RNA sequencing. Next to 133 significantly upregulated and 410 significantly downregulated genes in the ALS cohort compared to controls, the group identified the ubiquitin-proteasome pathway, the oxidative stress response, and the unfolded protein response to be upregulated based on Gene Ontology analysis^36^. Of these, qPCR was used for validation of two genes (ACTB and CUEDC2) where no significant changes were detected. The ubiquitin-proteasome pathway has been also found to be enriched in plasma derived EVs from another study that investigated the miRNA signature of EVs in ALS and additional neurodegenerative studies^47^.

A separate study that profiled miRNAs from plasma-derived EVs using next-generation sequencing also reported upregulation of the ubiquitin–proteasome pathway^39^. In EVs derived from persons with ALS, 5 miRNAs were upregulated whereas 22 miRNAs were downregulated compared to controls. Next to detected changes in the ubiquitin-proteasome pathway, in silico functional annotation of predicted mRNA targets of the identified deregulated miRNAs revealed affected transcriptional regulation process. The differentially expressed miRNAs were validated using qPCR in EVs isolated from 3 controls and 12 persons with ALS. Only one of the identified significantly deregulated miRNAs was validated with ddPCR (miR-15a-5p) which confirmed the significant upregulation identified with next-generation sequencing. Another study profiled miRNAs in plasma-derived EVs, distinguishing between putative smaller-sized EVs and larger-sized EVs using miRNA sequencing. The analysis revealed that the miRNA profile of EVs was distinct from blood cells, indicating selective packaging of ALS-relevant miRNAs within EVs^64^. In putative smaller-sized EVs 88 miRNAs were upregulated and 199 were downregulated whereas in putative larger-sized EVs 183 were upregulated and 215 were downregulated in persons with ALS compared to controls. They identified 8 miRNAs that were specifically found only in putative larger-sized EVs from persons with ALS and 22 miRNAs only in EVs from persons with ALS. While some of the differentially expressed miRNAs in putative larger-sized and smaller-sized EVs were used to perform pathway analyses, none of the identified mRNAs was validated with qPCR.

#### Diagnostic accuracy

To identify the best diagnostic biomarker(s) for ALS vs. controls, we pooled available studies using a BRMA model (**Figure 3A-B**). The model revealed a moderate diagnostic accuracy (AUC = 0.839; partial AUC accounting for FDR = 0.788) and heterogeneity (I² = 67.3%). As we reported previously for parkinsonian^18, 19^ and dementia^20^, RNA-associated biomarkers in general EVs appear to have the best combination of sensitivity and specificity (e.g., miR-126-5p). Publication bias assessment using Begg’s correlation (Kendall’s tau = 0.36, p = 0.12), Egger’s regression test (t = 1.82, p = 0.099) and visual inspection of funnel plot (**Figure 3C**) suggest no bias.

**Figure 3.**
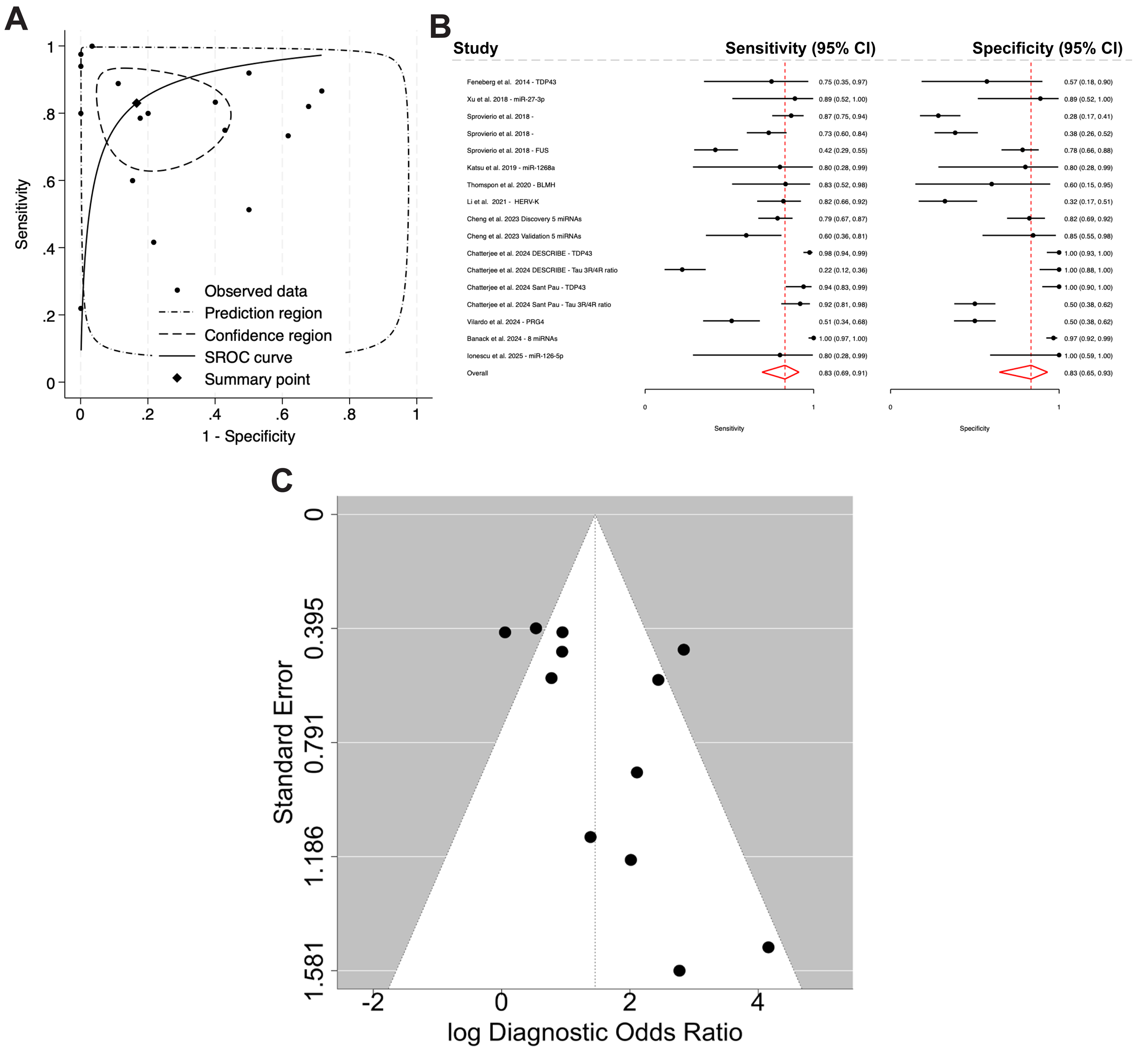
Diagnostic accuracy of general and CNS-enriched extracellular vesicle (EV) biomarkers for distinguishing ALS from controls. **(A)** Hierarchical summary receiver operating characteristic (HSROC) curve showing individual study estimates (dots), the summary point (diamond), 95% confidence region (dashed line), and 95% prediction region (dash-dot line). The solid curve represents the HSROC fit across included studies. **(B)** Forest plots of sensitivity and specificity for each biomarker study, grouped by analyte. Point estimates and 95% confidence intervals are shown for both sensitivity (left panel) and specificity (right panel), with red dashed lines indicating pooled summary estimates. **(C)** Funnel plot assessing publication bias for the included studies. Each point represents an individual study plotted by effect size (log diagnostic odds ratio) and standard error. Visual inspection shows no strong asymmetry, consistent with statistical tests indicating no evidence of publication bias.

## Discussion

In this study, we systematically reviewed the utility of EVs from 39 studies (**Table 1**) as a liquid biopsy for aiding in the diagnosis of ALS (**Figure 1**). Out of those studies, we meta-analyzed EV-associated TDP-43 levels in 5 studies using random-effects models and 11 in the diagnostic accuracy meta-analysis. The included studies spanned a wide range of EV isolation methods, biofluids and analytical platforms. Despite this extensive literature, our findings highlight substantial methodological heterogeneity, limited replication across research groups, and inconsistent biomarker performance.

Across all protein focused studies, the most extensively investigated analyte was EV associated TDP-43. Although majority of studies reported slightly higher TDP-43 levels in ALS, our pooled random effects meta-analysis demonstrated only a moderate SMD (**Figure 2A)** and very high heterogeneity without reaching statistical significance. A sensitivity analysis excluding one outlier study reporting unusually low values strengthened the overall effect, but heterogeneity remained high. Publication bias analyses using Begg correlation, Egger regression and funnel plot visualization (**Figure 2B**) showed minimal asymmetry, suggesting that the observed variability mainly reflects true methodological and biological differences between studies rather than selective reporting.

RNA based biomarkers were evaluated in 16 studies, ranging from targeted qPCR panels to RNA sequencing. In our two previous meta-analyses for parkinsonian disorders^18, 19^ and dementia^20^, RNA derived from general EVs consistently showed the highest diagnostic performance compared with other EV analytes. However, in ALS, the available studies displayed very poor overlap and often originated from the same research groups, which limits generalizability. But our diagnostic accuracy meta-analysis showed a moderate pooled performance with an AUC of approximately 0.84 and moderate heterogeneity (**Figure 3A-B**) with no publication bias (**Figure 3C**). However, these results remain preliminary because replication across independent cohorts is limited and validation using standardized isolation and quantification methods is lacking.

Although most studies to date have relied on either protein-based or RNA-based analyses of EV cargo, single-analyte biomarkers are unlikely to capture the full biological complexity of ALS. Given the multifactorial nature of the disease and the heterogeneous cellular processes reflected in EVs, future efforts will likely require integrated, multi-omics approaches^71^ that combine targeted protein, RNA, lipid, and metabolite profiling. Such composite biomarker panels may offer superior diagnostic accuracy, improve biological interpretability, and better reflect the diverse pathological pathways active in ALS.

A major source of inconsistency was the use of speculative CNS-enriched EVs. Multiple studies measuring miRNAs or proteins in neuronal or glial marker enriched EVs described differences between ALS and controls, but these studies were almost exclusively produced by the same research groups. None of the datasets were shared despite repeated requests, no independent replication exists, and the identified biomarkers had poor overlap with other studies. Moreover, prior meta-analyses have demonstrated that speculative CNS-enriched EVs perform markedly worse diagnostically and exhibit substantially greater publication bias compared with general EVs^18–20^. This is further compounded by ongoing uncertainty about their biological origin since the markers used for enrichment are not specific to neurons, exist in soluble form and EVs originating from various tissues undergo extensive circulation and uptake which likely dilutes any original signal from the CNS. Collectively, these factors raise significant concerns regarding their reliability, reproducibility, and validity as clinical biomarkers.

## Conclusion

Overall, the present synthesis highlights both the promise and the limitations of EV based biomarkers in ALS. General EVs appear more robust and reproducible than speculative CNS-enriched preparations and protein biomarkers such as TDP43 show biologically consistent but methodologically variable results. RNA signatures remain appealing due to their diagnostic performance in other neurodegenerative conditions, but ALS specific studies require significantly more replication and methodological standardization. Future investigations should prioritize transparent data sharing, harmonized EV isolation protocols, rigorous validation using orthogonal assays and replication across independent cohorts.

## Data Availability

All data produced in the present study are available upon reasonable request to the authors

## Sources of funding

None

## Acknowledgment

We thank all authors who responded to our request and provided the missing information.

## Conflict of interest

The authors declare no conflict of interest.

## Authorship contributions

HBT conceptualized and designed the study. HBT and MMB wrote the manuscript. HBT and MMB analyzed the data. NV and HBT created the tables. JS, AS, MMB, NV, FK, AZ, TR, SW, NGR, LW, GR, AB and HBT collected data and checked the article for accuracy.

## Notes

### Competing Interest Statement

The authors have declared no competing interest.

### Funding Statement

This study did not receive any funding

## References

1. Mackenzie, I.R. et al. Pathological TDP-43 distinguishes sporadic amyotrophic lateral sclerosis from amyotrophic lateral sclerosis with SOD1 mutations. Ann Neurol 61, 427–434 (2007).

2. Neumann, M. et al. Ubiquitinated TDP-43 in frontotemporal lobar degeneration and amyotrophic lateral sclerosis. Science 314, 130–133 (2006).

3. Al-Chalabi, A. & Hardiman, O. The epidemiology of ALS: a conspiracy of genes, environment and time. Nat Rev Neurol 9, 617–628 (2013).

4. Andersen, P.M. & Al-Chalabi, A. Clinical genetics of amyotrophic lateral sclerosis: what do we really know? Nat Rev Neurol 7, 603–615 (2011).

5. Richards, D., Morren, J.A. & Pioro, E.P. Time to diagnosis and factors affecting diagnostic delay in amyotrophic lateral sclerosis. J Neurol Sci 417, 117054 (2020).

6. Sennfalt, S. et al. The path to diagnosis in ALS: delay, referrals, alternate diagnoses, and clinical progression. Amyotroph Lateral Scler Frontotemporal Degener 24, 45–53 (2023).

7. Hardiman, O., et al. Amyotrophic lateral sclerosis. Nat Rev Dis Primers 3, 17071 (2017).

8. Staats, K.A., Borchelt, D.R., Tansey, M.G. & Wymer, J. Blood-based biomarkers of inflammation in amyotrophic lateral sclerosis. Mol Neurodegener 17, 11 (2022).

9. Vu, L.T. & Bowser, R. Fluid-Based Biomarkers for Amyotrophic Lateral Sclerosis. Neurotherapeutics 14, 119–134 (2017).

10. Mazon, M., Vazquez Costa, J.F., Ten-Esteve, A. & Marti-Bonmati, L. Imaging Biomarkers for the Diagnosis and Prognosis of Neurodegenerative Diseases. The Example of Amyotrophic Lateral Sclerosis. Front Neurosci 12, 784 (2018).

11. McMackin, R., Bede, P., Ingre, C., Malaspina, A. & Hardiman, O. Biomarkers in amyotrophic lateral sclerosis: current status and future prospects. Nat Rev Neurol 19, 754–768 (2023).

12. de Carvalho, M. et al. Electrodiagnostic criteria for diagnosis of ALS. Clin Neurophysiol 119, 497–503 (2008).

13. Goutman, S.A. et al. Recent advances in the diagnosis and prognosis of amyotrophic lateral sclerosis. Lancet Neurol 21, 480–493 (2022).

14. van Niel, G., D’Angelo, G. & Raposo, G. Shedding light on the cell biology of extracellular vesicles. Nat Rev Mol Cell Biol 19, 213–228 (2018).

15. Colombo, M., Raposo, G. & Thery, C. Biogenesis, secretion, and intercellular interactions of exosomes and other extracellular vesicles. Annu Rev Cell Dev Biol 30, 255–289 (2014).

16. Hill, A.F. Extracellular Vesicles and Neurodegenerative Diseases. J Neurosci 39, 9269–9273 (2019).

17. Dutta, S., Hornung, S., Taha, H.B. & Bitan, G. Biomarkers for parkinsonian disorders in CNS-originating EVs: promise and challenges. Acta Neuropathol 145, 515–540 (2023).

18. Taha, H.B. & Bogoniewski, A. Analysis of biomarkers in speculative CNS-enriched extracellular vesicles for parkinsonian disorders: a comprehensive systematic review and diagnostic meta-analysis. J Neurol 271, 1680–1706 (2024).

19. Taha, H.B. & Bogoniewski, A. Extracellular vesicles from bodily fluids for the accurate diagnosis of Parkinson’s disease and related disorders: A systematic review and diagnostic meta-analysis. J Extracell Biol 2, e121 (2023).

20. Taha, H.B. Alzheimer’s disease and related dementias diagnosis: a biomarkers meta-analysis of general and CNS extracellular vesicles. npj Dementia (2025).

21. Dellar, E.R. et al. Extracellular vesicles in TDP-43 proteinopathies: pathogenesis and biomarker potential. Mol Neurodegener 20, 68 (2025).

22. Chatterjee, M. et al. Plasma extracellular vesicle tau and TDP-43 as diagnostic biomarkers in FTD and ALS. Nat Med 30, 1771–1783 (2024).

23. Wan, X., Wang, W., Liu, J. & Tong, T. Estimating the sample mean and standard deviation from the sample size, median, range and/or interquartile range. BMC Med Res Methodol 14, 135 (2014).

24. Ruopp, M.D., Perkins, N.J., Whitcomb, B.W. & Schisterman, E.F. Youden Index and optimal cut-point estimated from observations affected by a lower limit of detection. Biom J 50, 419–430 (2008).

25. Harbord, R.M., Deeks, J.J., Egger, M., Whiting, P. & Sterne, J.A. A unification of models for meta-analysis of diagnostic accuracy studies. Biostatistics 8, 239–251 (2007).

26. Nyaga, V.N. & Arbyn, M. Metadta: a Stata command for meta-analysis and meta-regression of diagnostic test accuracy data - a tutorial. Arch Public Health 80, 95 (2022).

27. Nyaga, V.N. & Arbyn, M. Comparison and validation of metadta for meta-analysis of diagnostic test accuracy studies. Res Synth Methods 14, 544–562 (2023).

28. Zhou, Y. & Dendukuri, N. Statistics for quantifying heterogeneity in univariate and bivariate meta-analyses of binary data: the case of meta-analyses of diagnostic accuracy. Stat Med 33, 2701–2717 (2014).

29. Lin, L. & Chu, H. Quantifying publication bias in meta-analysis. Biometrics 74, 785–794 (2018).

30. Feneberg, E. et al. Limited role of free TDP-43 as a diagnostic tool in neurodegenerative diseases. Amyotroph Lateral Scler Frontotemporal Degener 15, 351–356 (2014).

31. Tomlinson, P.R. et al. Identification of distinct circulating exosomes in Parkinson’s disease. Ann Clin Transl Neurol 2, 353–361 (2015).

32. Ding, X. et al. Exposure to ALS-FTD-CSF generates TDP-43 aggregates in glioblastoma cells through exosomes and TNTs-like structure. Oncotarget 6, 24178–24191 (2015).

33. Iguchi, Y. et al. Exosome secretion is a key pathway for clearance of pathological TDP-43. Brain 139, 3187–3201 (2016).

34. Xu, Q. et al. Comparison of the extraction and determination of serum exosome and miRNA in serum and the detection of miR-27a-3p in serum exosome of ALS patients. Intractable Rare Dis Res 7, 13–18 (2018).

35. Sproviero, D. et al. Pathological Proteins Are Transported by Extracellular Vesicles of Sporadic Amyotrophic Lateral Sclerosis Patients. Front Neurosci 12, 487 (2018).

36. Otake, K., Kamiguchi, H. & Hirozane, Y. Identification of biomarkers for amyotrophic lateral sclerosis by comprehensive analysis of exosomal mRNAs in human cerebrospinal fluid. BMC Med Genomics 12, 7 (2019).

37. Katsu, M. et al. MicroRNA expression profiles of neuron-derived extracellular vesicles in plasma from patients with amyotrophic lateral sclerosis. Neurosci Lett 708, 134176 (2019).

38. Chen, Y., Xia, K., Chen, L. & Fan, D. Increased Interleukin-6 Levels in the Astrocyte-Derived Exosomes of Sporadic Amyotrophic Lateral Sclerosis Patients. Front Neurosci 13, 574 (2019).

39. Saucier, D. et al. Identification of a circulating miRNA signature in extracellular vesicles collected from amyotrophic lateral sclerosis patients. Brain Res 1708, 100–108 (2019).

40. Banack, S.A., Dunlop, R.A. & Cox, P.A. An miRNA fingerprint using neural-enriched extracellular vesicles from blood plasma: towards a biomarker for amyotrophic lateral sclerosis/motor neuron disease. Open Biol 10, 200116 (2020).

41. Vassileff, N. et al. Revealing the Proteome of Motor Cortex Derived Extracellular Vesicles Isolated from Amyotrophic Lateral Sclerosis Human Postmortem Tissues. Cells 9 (2020).

42. Morasso, C.F. et al. Raman spectroscopy reveals biochemical differences in plasma derived extracellular vesicles from sporadic Amyotrophic Lateral Sclerosis patients. Nanomedicine 29, 102249 (2020).

43. Yelick, J. et al. Elevated exosomal secretion of miR-124-3p from spinal neurons positively associates with disease severity in ALS. Exp Neurol 333, 113414 (2020).

44. Chen, P.C. et al. Exosomal TAR DNA-binding protein-43 and neurofilaments in plasma of amyotrophic lateral sclerosis patients: A longitudinal follow-up study. J Neurol Sci 418, 117070 (2020).

45. Thompson, A.G. et al. CSF extracellular vesicle proteomics demonstrates altered protein homeostasis in amyotrophic lateral sclerosis. Clin Proteomics 17, 31 (2020).

46. Hayashi, N. et al. Proteomic analysis of exosome-enriched fractions derived from cerebrospinal fluid of amyotrophic lateral sclerosis patients. Neurosci Res 160, 43–49 (2020).

47. Sproviero, D. et al. Different miRNA Profiles in Plasma Derived Small and Large Extracellular Vesicles from Patients with Neurodegenerative Diseases. Int J Mol Sci 22 (2021).

48. Li, Y., Chen, Y., Zhang, N. & Fan, D. Human endogenous retrovirus K (HERV-K) env in neuronal extracellular vesicles: a new biomarker of motor neuron disease. Amyotroph Lateral Scler Frontotemporal Degener 23, 100–107 (2022).

49. Pasetto, L. et al. Decoding distinctive features of plasma extracellular vesicles in amyotrophic lateral sclerosis. Mol Neurodegener 16, 52 (2021).

50. Pregnolato, F. et al. Exosome microRNAs in Amyotrophic Lateral Sclerosis: A Pilot Study. Biomolecules 11 (2021).

51. Lo, T.W. et al. Extracellular Vesicles in Serum and Central Nervous System Tissues Contain microRNA Signatures in Sporadic Amyotrophic Lateral Sclerosis. Front Mol Neurosci 14, 739016 (2021).

52. Sproviero, D. et al. Extracellular Vesicles Derived From Plasma of Patients With Neurodegenerative Disease Have Common Transcriptomic Profiling. Front Aging Neurosci 14, 785741 (2022).

53. Sjoqvist, S. & Otake, K. A pilot study using proximity extension assay of cerebrospinal fluid and its extracellular vesicles identifies novel amyotrophic lateral sclerosis biomarker candidates. Biochem Biophys Res Commun 613, 166–173 (2022).

54. Zhou, Q. et al. Increased expression of coronin-1a in amyotrophic lateral sclerosis: a potential diagnostic biomarker and therapeutic target. Front Med 16, 723–735 (2022).

55. Banack, S.A., Dunlop, R.A., Stommel, E.W., Mehta, P. & Cox, P.A. miRNA extracted from extracellular vesicles is a robust biomarker of amyotrophic lateral sclerosis. J Neurol Sci 442, 120396 (2022).

56. Salomon-Zimri, S. et al. Combination of ciprofloxacin/celecoxib as a novel therapeutic strategy for ALS. Amyotroph Lateral Scler Frontotemporal Degener 24, 263–271 (2023).

57. Cheng, Y.F. et al. Signature of miRNAs derived from the circulating exosomes of patients with amyotrophic lateral sclerosis. Front Aging Neurosci 15, 1106497 (2023).

58. Sjoqvist, S. & Otake, K. Saliva and Saliva Extracellular Vesicles for Biomarker Candidate Identification-Assay Development and Pilot Study in Amyotrophic Lateral Sclerosis. Int J Mol Sci 24 (2023).

59. Wajnberg, G. et al. Application of annotation-agnostic RNA sequencing data analysis tools for biomarker discovery in liquid biopsy. Front Bioinform 3, 1127661 (2023).

60. Vilardo, B. et al. Shotgun Proteomics Links Proteoglycan-4(+) Extracellular Vesicles to Cognitive Protection in Amyotrophic Lateral Sclerosis. Biomolecules 14 (2024).

61. Banack, S.A. et al. A microRNA diagnostic biomarker for amyotrophic lateral sclerosis. Brain Commun 6, fcae268 (2024).

62. Raineri, D. et al. Circulating GLAST(+) EVs are increased in amyotrophic lateral sclerosis. Front Mol Biosci 11, 1507498 (2024).

63. Al Ojaimi, Y. et al. Metabolomic and Proteomic Profiling of Serum-Derived Extracellular Vesicles from Early-Stage Amyotrophic Lateral Sclerosis Patients. J Mol Neurosci 75, 21 (2025).

64. Dragoni, F. et al. Cross-tissue MiRNA profiling of extracellular vesicles and PBMCs from amyotrophic lateral sclerosis patients. Sci Rep 15, 14976 (2025).

65. Gautam, M. et al. Exosome Proteomics of SOD1(D90A) Mutation Suggest Early Disease Mechanisms, and FN1 as a Biomarker. Ann Clin Transl Neurol (2025).

66. Ionescu, A. et al. Muscle-derived miR-126 regulates TDP-43 axonal local synthesis and NMJ integrity in ALS models. Nat Neurosci 28, 2201–2216 (2025).

67. Kato, C. et al. Proteomic insights into extracellular vesicles in ALS for therapeutic potential of Ropinirole and biomarker discovery. Inflamm Regen 44, 32 (2024).

68. Mead, R.J., Shan, N., Reiser, H.J., Marshall, F. & Shaw, P.J. Amyotrophic lateral sclerosis: a neurodegenerative disorder poised for successful therapeutic translation. Nat Rev Drug Discov 22, 185–212 (2023).

69. Gaetani, L. et al. Neurofilament light chain as a biomarker in neurological disorders. J Neurol Neurosurg Psychiatry 90, 870–881 (2019).

70. Chio, A. et al. Phenotypic heterogeneity of amyotrophic lateral sclerosis: a population based study. J Neurol Neurosurg Psychiatry 82, 740–746 (2011).

71. Li, S. et al. Advancing biological understanding of cellular senescence with computational multiomics. Nat Genet 57, 2381–2394 (2025).

