## Supplementary material for "Extracellular vesicles as a liquid biopsy for amyotrophic lateral sclerosis: a systematic review and meta-analysis": Table S1

**Table S1.** Complete search strategy using PUBMED and EMBASE. The search was performed from the date of inception until October 25th, 2025.

| | |
|---|---|
| PUBMED | (Extracellular vesicles[Title/Abstract] OR Extracellular vesicle[Title/Abstract] OR Exosomes[Title/Abstract] OR Exosome[Title/Abstract] OR "Apoptotic body"[Title/Abstract] OR "Apoptotic Bodies"[Title/Abstract] OR "EV"[Title/Abstract] OR "EVs"[Title/Abstract] OR Ectosome[Title/Abstract] OR Ectosomes[Title/Abstract] OR "Microvesicle"[Title/Abstract]) AND ("amyotrophic lateral sclerosis"[Title/Abstract] OR "ALS"[Title/Abstract] OR "motor neuron disease"[Title/Abstract] OR "MND"[Title/Abstract] OR "Lou Gehrig's disease"[Title/Abstract] OR "classical ALS"[Title/Abstract] OR "sporadic ALS"[Title/Abstract] OR "familial ALS"[Title/Abstract] OR "bulbar-onset ALS"[Title/Abstract] OR "bulbar onset ALS"[Title/Abstract] OR "limb-onset ALS"[Title/Abstract] OR "primary lateral sclerosis"[Title/Abstract] OR "PLS"[Title/Abstract] OR "progressive muscular atrophy"[Title/Abstract] OR "PMA"[Title/Abstract] OR "ALS-FTD"[Title/Abstract]) NOT 'Review'[Publication Type] |
| EMBASE | ('extracellular vesicle':ti,ab OR 'extracellular vesicles':ti,ab OR exosome:ti,ab OR exosomes:ti,ab OR 'apoptotic body':ti,ab OR 'apoptotic bodies':ti,ab OR ev:ti,ab OR evs:ti,ab OR ectosome:ti,ab OR ectosomes:ti,ab OR microvesicle:ti,ab) AND ('amyotrophic lateral sclerosis':ti,ab OR als:ti,ab OR 'motor neuron disease':ti,ab OR mnd:ti,ab OR 'lou gehrig* disease':ti,ab OR 'classical als':ti,ab OR 'sporadic als':ti,ab OR 'familial als':ti,ab OR 'bulbar-onset als':ti,ab OR 'bulbar onset als':ti,ab OR 'limb-onset |

| | |
|---|---|
| | als':ti,ab OR 'primary lateral sclerosis':ti,ab OR pls:ti,ab OR 'progressive muscular atrophy':ti,ab OR pma:ti,ab OR 'als-ftd':ti,ab) NOT 'review'/it |

**Table S2.** Differentially expressed proteins from proteomics and proximity extensions assay studies.

| Study, Country, PMID | Protein(s) | Direction | FC/Effect Size | EV type; notes |
|---|---|---|---|---|
| Feneberg et al. 2014 Germany 24834468 | TDP-43 | NS | NS | CSF EVs |
| Tomilson et al. UK 2015 25909081 | PGK1; GAPDH; GNB1; ACTB; CCL5; FGB; RAP1B; C1QB; HLA-A; C1QA; C1QC; PON1; S100A9; S100A8; AHSG; ITGB1; UBR4; KIF14; CD36; FERMT3; UBA52; SLC2A1; AGT; DCD; VPS13D; SDCBP; RSU1; PRSS2; CFL1; PRDX2; F2; CYBB; COL1A2; HSPA7; TLL1; LPA | ↓ ALS vs. PD | Infinity; 3.41; 8.42; 3.93; 14.99; 2.29; 9.31; 1.69; 5.29; 1.65; 1.63; 2.58; 3.22; 7.35; 3.85; 4.42; infinity; 1.71; 3.94; 8.97; 1.69; 2.7; 2.09; 5.94; 8.46; 2.43; 3.58; 2; 3.67; 9.57; 1.68; 1.87; 1.99; 1.92; ; 3.73; 2.84 | Serum EVs |
| | C8G; IGHV3; C4BPA; PROS1; CPAMD8; IGLV1; A2M; IGKV1; IGKV3; C4A; ADIPOQ; CP; C4BPB; TNRC18; IGKV3. | ↑ ALS vs. PD | 5.17; 1.62; 1.42; 5.7; 3.51; 2.85; 2.46; 2.05; 1.61; 2.72; 1.6; 1.51; 2.08; 1.46 | Serum EVs |
| Vassileff et al. 2020 Australia 32708779 | CD177, CHMP4B, CSPG5, DYNC1I2, IGHV3-43, LBP, RPS29, S100A9, SAA1, SCAMP4, SCN2B, SLC16A1 | Present only in ALS EVs | NR | Cortex EVs |
| | VCAM1, RRAS, PLSCR4, NT5E, ITGA5, HLA-A, GYPC, ENPP6, ENG, | ↓ ALS vs. controls | NR | Cortex EVs |

| | | | | |
|---|---|---|---|---|
| | EHD1, BST1, AHNAK | | | |
| | STAU1, FXYD6, DYNC1I1, DHX30 | ↑ ALS vs. controls | NR | Cortex EVs |
| Thompson et al. 2020 UK 32821252 | BLMH | ↓ ALS vs. controls | NR | CSF EVs |
| | Proteasome Core Complex (Functional Group: PSMA4, PSMA5, PSMA6, PSMA7, PSMB1, PSMB2, PSMB3, PSMB6) | ↓ ALS vs. controls | NR | Gene Set Enrichment CSF EVs; Analysis revealed a significant diminution of this functional pathway. All eight proteins annotated to this complex showed a negative log fold change, indicating a general decrease in proteasomal components |
| | UBA1, ANXA11, CYBB, CFL1 | ↓ C9orf72 ALS vs. non-C9orf72 ALS | NR | CSF EVs |
| | GPNMB, TGM2 | ↑ C9orf72 ALS vs. non-C9orf72 ALS | NR | CSF EVs |
| Hayashi et al. 2020 Japan 31669371 | NOC2L, PDCD6IP, VCAN | ↑ ALS vs. iNPH | 2.39; 1.39; 1.36 | CSF EVs |
| | SERPINA3, PTPRZ1, C1QC, CCDC19, MYL6B, MARCO, FCGBP, FOLR1, RELN, CFB, CHMP4A | ↓ ALS vs. iNPH | 3.73; 2.49; 2.05; 3.02; 2.37; 1.70; 1.77; 2.08; 2.45; 2.27; 1.84 | CSF EVs |
| Zhou et al. 2022 China 35648369 | HNRNPD, HNRNPA0, HNRNPC, HNRNPA1, RBMX, PUF60, TCERG1, SF3B3, DEK, DDX5, CORO1A, ACTG1, ACTG2, ARPC1B, IQGAP1, FUS, PADI4 | ↑ ALS vs. controls | NR | Plasma EVs |

| | | | | |
|---|---|---|---|---|
| Kato et al. Japan 2024 38997748 | SERPING1, C4A, C4B, C5, C1RL, COLEC10, FRGP1, PROS1, SERPINA5, SERPINA10, SERPINA12, SERPINI, SERPIND1 (and others) | ↑ALS vs. controls | NR | Identified as commonly elevated (31 DAPs total) in both serum EVs (sEVs) and CSF EVs (cEVs) of SALS patients (ROPI-naive) S to controls, associated with Complement/Coagulation Cascades |
| | HSP90AA1, HSPA1A, HSPA14, MAP2K2, TXT, ANXA3, ARGC1, ARPC4, CFL1, PFN | ↓ ALS vs. controls | NR | Identified as commonly decreased (38 DAPs total) in both sEVs and cEVs of SALS patients (ROPI-naive) compared to controls, associated with Unfolded Protein Response (UPR) and actin regulation. |
| Ojaimi et al. 2025 France 39954028 | TALDO1, PTRC, PIP4K2A, LGALS7, GP9, TMP3, FGB, P4HB, CD36 | ↑ALS vs. controls | NR | Serum EVs |
| | SERPING1, IGHA2, IGLC3, APOC1, APOB, F11, SLC4A1, APMAP | ↓ ALS vs. controls | NR | Serum EVs |
| Vilardo et al. 2024 Italy 38927130 | PRG-4, LBP, VWF, FIBA, FIBB, FIBG, C09 | ↑ ALS vs. controls | 1.72, 1.84, 1.58, 1.36, 1.41, 1.45, 1.33 (Discovery FCs); 2.07, 1.56, 1.75, 1.75, 1.58, 1.50, 1.41 (Validation FCs) | Plasma EVs |
| | LEG3, TGM3, PLAK, FBLN3, DESP, TRY1, | ↓ ALS vs. controls | 0.11; 0.25; 0.21; 0.33; 0.29; 0.39; | Plasma EVs |

| | | | | |
|---|---|---|---|---|
| | APOD | | 0.64 | |
| Gautam et al. 2025 USA 41024384 | FN1 | ↑ in more advanced ALS | NR | Serum EVs |
| Sjoqvist et al. 2023 Japan 36982312 | None | None | None | Saliva EVs |
| Sjoqvist et al. Japan 2022 35567903 | None | None | None | CSF EVs |
